# The natural history of the emergence of sexually transmissible shigellosis

**DOI:** 10.1101/2025.05.02.25326874

**Authors:** Lewis C. E. Mason, Fariha Jawed, Angelika Fruth, Roberto Vivancos, Claire Jenkins, Kate S. Baker

## Abstract

Shigellosis is a gastrointestinal illness caused by bacteria belonging to one of four species of *Shigella*. Sexually transmissible shigellosis was first reported in 1974, but recently there has been a global increase in the transmission of extensively drug-resistant strains. Here, we sought to characterise the natural history of sexually transmissible shigellosis through literature review and genomic epidemiological analysis of early outbreaks. The literature review revealed a significant gap in reporting of sexually transmissible shigellosis between the first report in 1974 and the early 2000s after which reporting increased. To better understand this sustained emergence of sexually transmissible shigellosis in the 21^st^ century, we explored potential pathogen factors and linked these with changes in host populations. Specifically, we analysed the genomic epidemiology of preserved strains from outbreaks in both Berlin (2000 – 2002) and London (2004 – 2006). Both outbreaks were *S. sonnei* Genotype 3.1, an ancestral branch of the globally disseminated Lineage III subtype, which is distinct from the currently globally dominant extensively drug-resistant forms (Genotypes 3.6.1.1.2 and 3.6.1.1) circulating in sexual transmission networks. We also describe the variable antimicrobial resistance, conserved colicin genes, and differing virulence and plasmid profiles between the London and Berlin outbreaks. Finally, we conducted temporal reconstruction of Genotype 3.1 and found that the most recent common ancestor occurred in 1999 (95% HPD 1998 – 2000) which is coincident with the introduction of highly active antiretroviral therapy (HAART) for HIV. This suggests that changes associated with the introduction of HAART likely contributed to the re-emergence of sexually transmissible shigellosis in the 21^st^ Century.

**Data Summary:** Genome accession numbers as associated isolate metadata are available in the Supplementary Data Table.

**Impact Statement:** This work explores the natural history of the sustained re-emergence of sexually transmissible (ST) shigellosis in the 21^st^ Century. Insights from our analysis of the genomic epidemiology of early ST *S. sonnei* outbreaks in London and Berlin highlighted the importance of culturing and sequencing historical bacterial pathogenic isolates. We demonstrated the importance of AMR and plasmid dynamics as drivers of ST outbreaks, as the associated 3.1 Genotype of *S. sonnei* was later replaced by Genotypes 3.6.1.1.2 and 3.6.1.1. The latter contemporarily dominates the landscape of extensively drug-resistant (XDR) *S. sonnei* driven by the stepwise acquisition of azithromycin, ciprofloxacin, and ceftriaxone resistance determinants. We further highlight the role that evolutionary refinement in other parts of the bacterial accessory genome (e.g. colicins, virulence genes) may contribute to the successful proliferation, transmission and persistence of *S. sonnei*. Finally, we provide temporal evidence that the availability of HIV highly activate antiretroviral therapy (HAART) may have been a factor in driving the emergence of endemic sexually transmissible shigellosis.

## Introduction

*Shigella* is a genus of bacteria which causes shigellosis, a severe gastrointestinal illness. Shigellosis is characterised by a range of various common, and rarer, more serious symptoms including bloody diarrhoea (dysentery), vomiting, stomach cramps, rectal prolapse, dehydration, reactive arthritis, and toxic megacolon (1). Traditionally, *S. sonnei* outbreaks have been associated with travel to low-and middle-income countries (LMICs) and consumption of faeces-contaminated food and water (2).

However, in 1974 (3), an outbreak of sexually transmissible (ST) shigellosis was reported occurring through faecal-oral contact among gay, bi, and other men who have sex with men (GBMSM). Following this initial report, sporadic urban outbreaks were reported (e.g. in Berlin (4), London (5), San Francisco (6), and New South Wales, Australia (7)). In 2015, a large international genomic epidemiology study revealed the global linkage of contemporaneous ST shigellosis outbreaks (8).

Recent surveillance data has revealed that ST shigellosis is now endemic in many regions globally. Contemporary shigellosis outbreaks in GBMSM are characterised as being highly antimicrobial resistant, with XDR isolates resistant to azithromycin, ciprofloxacin, and ceftriaxone emerging globally (9) (10) (11) (12). This is mediated through the acquisition of AMR gene-harbouring plasmids (13), and the development of quinolone resistance determining region (QRDR) mutations found in the chromosomal DNA (14). Treatment options in GBMSM populations are limited due to increasing resistance to three out of the four WHO-recommended antimicrobials in the treatment of shigellosis (15) (16). Understanding the global spread of this threat to public health is critical to future control efforts, and may provide valuable learnings for other sexually transmissible enteric illnesses (e.g. *Campylobacter*) (17).

For this reason, we sought to better characterise the natural history of sexually transmissible shigellosis through a semi-systematic literature review of outbreak reporting since 1974 and a genomic epidemiology study of two of the earliest 21^st^ Century outbreaks of ST *S. sonnei.* Specifically, analysis of 23 isolates from outbreaks reported in 2004 - 2006 in London and 2000 - 2002 in Berlin (4) (5). In doing so, we reveal a quiescence in outbreak reporting between 1974 and the early 2000s, and find potential pathogen and host drivers for the emergence of ST shigellosis in the 21^st^ Century.

### Methods Literature review

To obtain an overview of changes in the reports of sexually transmissible shigellosis throughout time and space, we undertook a literature review using the following search terms in Scopus at 11:00 (GMT) on 30 January 2024: *(TITLE-ABS-KEY (shigell*) AND TITLE-ABS-KEY (gay) OR TITLE-ABS KEY (homosexual) OR TITLE-ABS KEY (men AND who AND have AND sex AND with AND men) OR TITLE-ABS KEY (msm) OR TITLE-ABS-KEY (gay AND bisexual AND other AND men-who-have-sex-with-men) OR TITLE-ABS-KEY (gbmsm) AND TITLE-ABS-KEY (outbreak) OR TITLE-ABS-KEY (cluster) OR TITLE-ABS-KEY (trends) OR TITLE-ABS-KEY (venue))*.

All articles (*n*=89) identified by Scopus were screened by two reviewers reading the full article. Inclusion criteria were: primary research articles reporting on at least one case of sexually transmissible shigellosis in GBMSM (or equivalent terminology). Articles that were reviews, or which did not explicitly state the sexually transmissible nature of disease were excluded. Description of ‘domestically acquired’ shigellosis alone was not sufficient to fulfil the definition of sexually transmissible shigellosis. The cut-off date for articles included in 2024 was 30 January 2024. Research articles visualised on the world map were identified using Scopus using identical search terms to those described above. Two individuals (LCEM, FJ) initially reviewed articles for inclusion/exclusion according to the criteria above, and discrepancies (*n* = 16/89) were resolved by a third reviewer blinded to initial scoring (KSB). Specific data (country, year of publication, and references to isolates being ciprofloxacin or azithromycin resistant) were then extracted and analysed from articles. In synthesis here, ‘outbreak reporting’ means the raw number of *Shigella* spp. outbreaks published in a given year, including instances where a country has reported multiple outbreaks. ‘Novel regional outbreak reporting’ refers to the number of different countries reporting shigellosis outbreaks.

## Genomic epidemiology study

### Data collection

#### Whole-genome sequenced isolates

Sequence data from outbreak isolates comprised Illumina Whole Genome Sequencing (WGS) of the London 2004 – 2006 outbreak isolates (*n*=14, from (5)) undertaken by the UK Health Security Agency (UKHSA) (Public Health England at the time) using a previously described methodology (18). Data from isolates in the Berlin 2000 – 2002 outbreak (*n*=9, from (4)) were Illumina sequenced as per in-house procedures at the Robert Koch Institute, as previously described using Illumina MiSeq with the Nextera XT DNA library kit (19).

Data for globally representative contextual isolates of *S. sonnei* subclades were also included in genomic and phylogenetic analyses via access through the Sequence Read Archive (20). Of the 120 reference isolates stipulated in previous work in the creation of a global genotyping framework (21), 113 were included (94%, *n* = 113/120) in this study, with six being omitted due to their unavailability for download from the SRA database, and one being omitted due to failing quality control analyses (having 75% missing data for assembly). All individual isolate accessions, associated metadata, and genomic analysis results can be found in the **Supplementary Data Table**.

#### Genomic analyses

The FASTQ files for the London outbreak and reference isolates used in this study were downloaded from the SRA (20), using *fastq-dump* SRA toolkit (v 2.11.0) (20). Trimmomatic (v 0.39) (22) was used to trim the sequences, with the following parameters: *2:30:10 LEADING: 20 TRAILING: 20 SLIDINGWINDOW: 4: 20 MINLEN: 40*. The trimmed reads were then checked for quality by using fastQC (v 0.11.9) (23), the results of which were combined with multiQC (v 1.12) (24).

To generate a multiple sequence alignment for phylogenetic inference, *S. sonnei* 53G (GenBank: *GCA_00283715.1_ASM28371v1*) was used as a reference genome. BWA (v 0.7.17) (25) was used to map the experimental genomes with the reference genome, where PCR duplicates were then removed with PICARD (v 2.27.2) (26). Qualimap (v 2.2.2) (27) bamqc was used to check the quality of the mapping.

#### Phylogenetic tree creation and visualisation

SAMtools (v 1.11) (28) and BCFtools (v 1.9) were used for variant calling. SNP-sites - C (v. 2.5.1) (29) was used on the core alignment multi-FASTA file to determine the invariant sites (IS). Using the parameters: *-fconst [IS1],[IS2],[IS3],[IS4]-keep-ident - bb 1000-m GTR+F+I+G4*, the phylogenetic trees were created using IQtree (v 2.2.0.3) (30) with the FASTA alignment output of filtered polymorphic sites generated by Gubbins (v 3.2.1) (31). The tree output from IQTree was then visualised in iTOL (32).

### Assembly

The genomes of all isolates were assembled using UNICYCLER (v 0.5.0) (33), with the trimmed forward/reverse paired/unpaired reads, using default parameters. The quality of these assemblies was checked using QUAST (v 5.0.2) (34).

### Gene and plasmid detection

AMR, stress, and virulence genes were identified using NCBI AMRFinder Plus (v 3.10.24) (35), with the assembled genomes of each isolate and the following parameters:--nucleotide assembly.fasta--organism *Escherichia*--plus. The presence and absence of colicin genes in the genome assemblies were investigated, using a previously generated custom database (36), embedded in ABRicate (v. 1.0.1), upon draft assemblies as previously described (37) using default parameters. The presence and absence of virulence genes were further investigated using VFDB (38), embedded in Abricate (v.1.0.1). Plasmids were detected using the PlasmidFinder database (39), embedded in ABRicate (v. 1.0.1). Mapping of the isolates to pKSR100 (GenBank accession CP090162) and pAPR100 (GenBank accession CP090161) using each as a reference (as in genomic analyses section above), then Qualimap (v 2.2.2) (27) was used to determine the 20X depth average mapping coverage (%) of the isolates to each of the two plasmids. SonneiTyping Script (v 20210201) (21) embedded in Mykrobe (v 0.11.0) (40) was used for genotyping and in the identification of QRDR mutations in the *gyrA* and *parC* genes.

### Temporal signal and dating analyses

A phylogenetic tree, containing solely the London and Berlin isolates, was generated using the methodology described above. The resulting tree file was then imported into TempEst (v 1.5.3) (41), alongside the exact dates of isolation (dd/mm/yyyy) for all isolates. The temporal signal was visualised with the best-fitting root and heuristic residual mean squared function. Dates were specified with the *’since some time in the past’* function. The resulting adequate temporal signal informed our decision to use the more resource-intensive Bayesian Evolutionary Analyses by Sampling of Trees (BEAST, v 2.7.7) package (42).

For BEAST analysis, the filtered polymorphic site sequence alignment output of the London and Berlin outbreak isolates was imported into BEAUti (v 2.7.7). The exact tip dates of the isolates were specified’as dates with format: dd/mm/yyyy’. The Gamma Site Model (Category Count: 8), GTR substitution model, Strict Clock model, coalescent constant population, and a Markov Chain Monte Carlo (MCMC) chain length of 10 million were used. The results of the three chains were combined using LogCombiner (v 2.7.7), using default parameters (burn in 10%, subsampling not used). The combined log was then visualised in Tracer (v 1.7.2) and evaluated for convergence based on the criterion: Effective Sampling Size (ESS) > 200 (43), where our’Tree.height’ analysis produced an ESS of 6356. The’Tree.height’ trace mean value was then used to infer the number of years (to four decimal places) ago the most recent common ancestor (MRCA) between the London and Berlin isolates emerged (from the exact date of the most recent isolate in the tree).

## Results and Discussion

### An overview of sexually transmissible *Shigella* spp. outbreak reporting

As a proxy for the epidemiological surveillance of sexually transmissible shigellosis, we examined the scientific literature for trends in reporting over time. Specifically, a structured literature review identified 66 articles reporting cases and outbreaks of *Shigella* infection among GBMSM (**Figure 1, Supplementary Data Table**) published since the first reported outbreak in 1974 (44). After reports of the early outbreaks in the 1970s, a period of dormancy followed until 2001, with the literature search identifying only one further outbreak reported in 1977 (44). Subsequently, in the early 2000s, reports of ST shigellosis outbreaks increased and became more widely reported geographically (**Figures 2 and 3**). The number of ST outbreaks reported has changed over the years, with an initial increase in the early 2000s being followed by an acceleration in reporting from 2015 onward (**Figure 2A**), which coincided with an increase in reporting of resistance against key classes of antimicrobials (**Figure 2B**). Parallel to the increase in the number of outbreaks reported, there was also an increase in the geographical spread of reporting (**Figures 2A and 3**).

**Figure 1.**
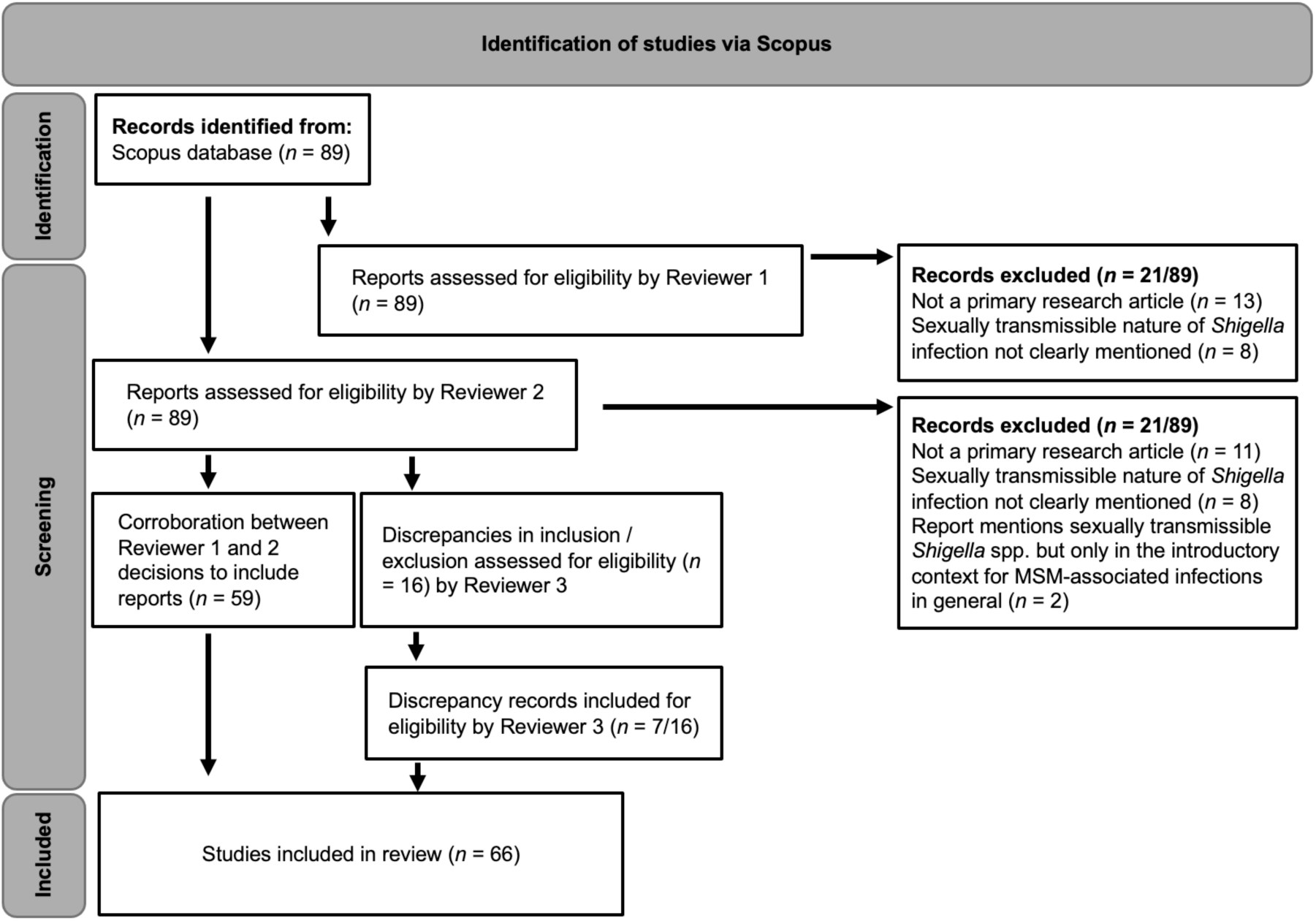
A flowchart showing the process of article screening during literature review.

**Figure 2.**
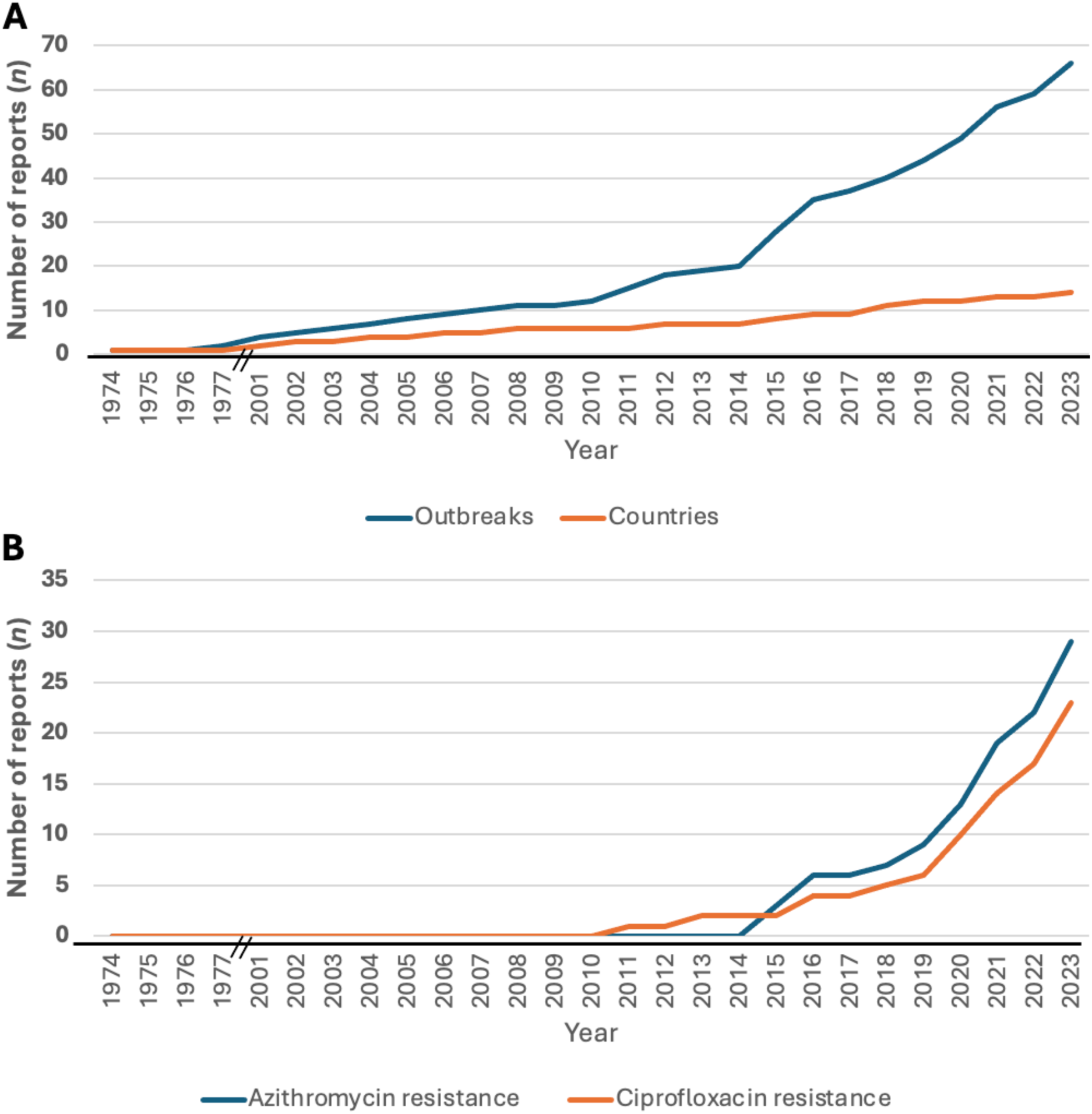
Trends in sexually transmissible shigellosis reporting. **(A)** Cumulative outbreak reporting of sexually transmissible shigellosis between 1974 – 2023 of counts of outbreaks (primary axis) and new countries reporting outbreaks (secondary axis). **(B)** Cumulative reporting of sexually transmissible shigellosis resistant to azithromycin and/or ciprofloxacin. Two black lines overlaying the X-axis represent the exclusion of years (1978 – 2000) owing to no reports of ST shigellosis being identified in the literature search.

**Figure 3.**
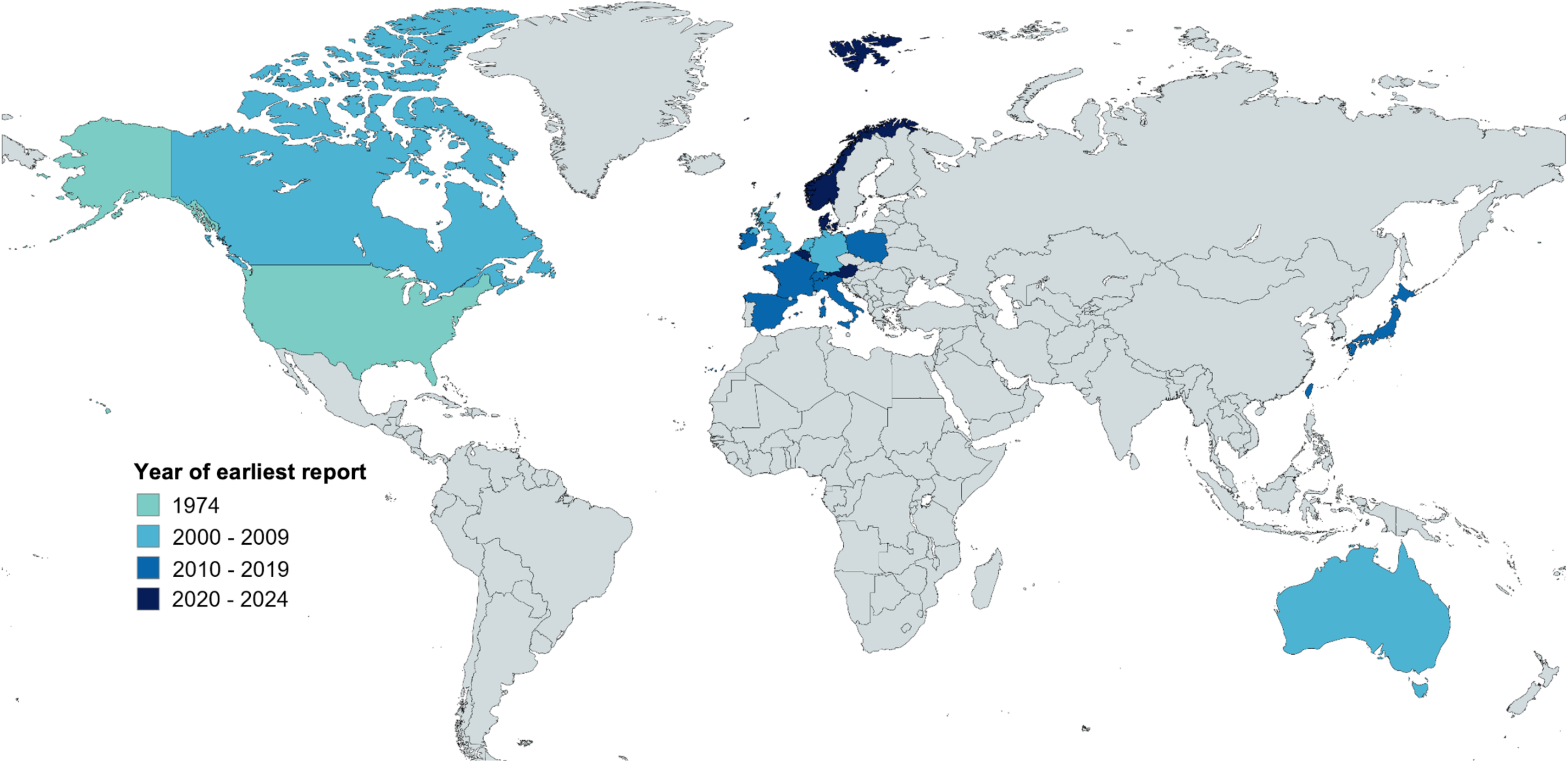
World map showing year of first reporting of ST shigellosis by location. The colour of the year (or year range) of the earliest publication date arising from the location is shown according to the inlaid key. The inclusion of Norway, Denmark, and Austria was via a recent European Centre for Disease Prevention and Control (ECDC) report (48) (not identified by literature review).

Although the increase in the number of outbreaks and countries reporting ST shigellosis likely represent genuine increased transmission, it is important to consider other factors that may have influenced these trends. There may have been underreporting attributable to the literature search terms (where e.g. ‘AND outbreak’ may have excluded case reports) and underreporting may also have occurred between 1974 and the early 2000s while the sexual health community focused on tackling the HIV pandemic. However, several factors point to reporting trends representing genuine increases in transmission including the known role of new technologies (e.g. geospatial phone applications) in driving ST shigellosis outbreaks (45), the association of outbreaks with AMR acquisition (8, 14, 46), and reports of increases over time throughout periods of consistent surveillance practices (47). Hence the reporting indicates an initial emergence in the early 1970s, following quiescence until the early 2000s, and then onward global spread accelerated through the rapid acquisition of antimicrobial resistance.

### Genomic epidemiology of early 21^st^ century ST *S. sonnei*

To investigate the relationships among the spate of sexually transmitting outbreaks of *S. sonnei* from the early 2000s, we contacted those reporting outbreaks from this period for a genomic epidemiology study. Two sites had retained their isolates and we proceeded with whole genome sequencing of *S. sonnei* isolates from outbreaks in: Berlin (*n*=9, sampled between 2000 and 2002, (4)) and London (*n*=14, sampled between 2004 – 2006 (5). We then constructed a phylogenetic tree of these isolates alongside representatives of the *S. sonnei* global genotyping framework (21) and genotyped the outbreak isolates. This revealed that, as anticipated, the early 2000s isolates were all (*n* = 23/23) the same genomic subtype (**Figure 4**). Namely Genotype 3.1, which is distinct from the currently globally predominant 3.6.1.1.2 and 3.6.1.1 sexually transmitting and extensively drug resistant Genotypes (9) (49) (50). Notably however, although the London and Berlin isolates were of the same Genotype, they fell into distinct phylogenetic clusters by city of origin, with a modest average pairwise distance (46 SNPs).

**Figure 4.**
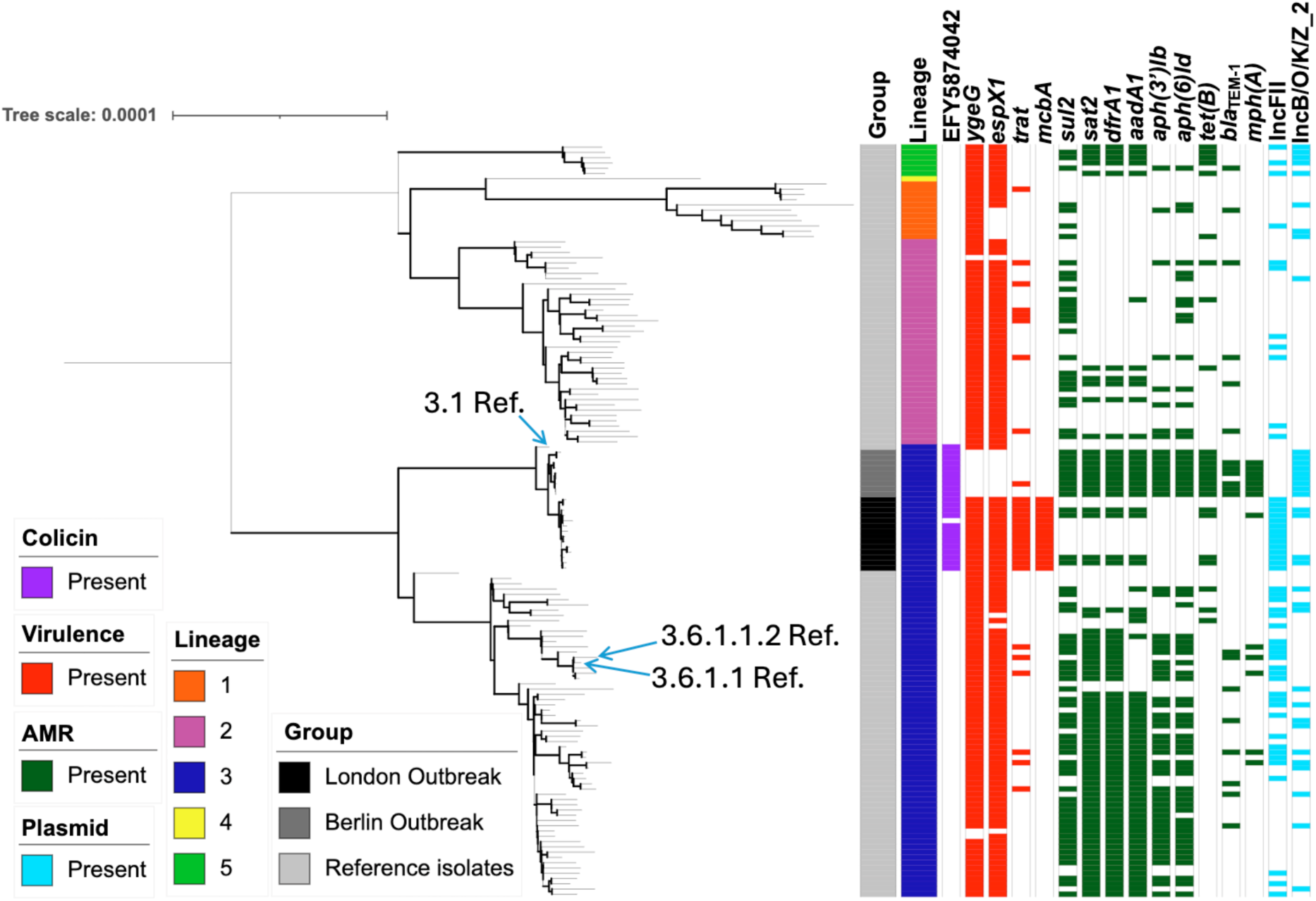
Phylogenetic relationships and accessory genome content among the London and Berlin outbreak, and contextual, *S. sonnei* isolates. A midpoint rooted maximum likelihood phylogenetic tree showing the associations between London outbreak isolates (*n* = 14), Berlin outbreak isolates (*n* = 9), and reference (Ref.) isolates (*n* = 113). Metadata tracks show, isolate group, Lineage, and the presence or absence of selected colicin, virulence, antimicrobial resistance (AMR) genes, and plasmid types, coloured according to the inlaid keys. Arrows adjacent to the tree indicate locations of the 3.1, 3.6.1.1, and 3.6.1.1.2 reference isolates in the tree. Emboldened branches are supported by bootstrap values of ≥ 70 of 100. Branch lengths represent substitutions per site across a 13,581 bp alignment.

While this analysis indicates that the outbreaks of the early 2000s may have been of a common globally disseminated Genotype, we were limited in the isolates that could be recovered to confirm this hypothesis. We had also reached out to corresponding authors (or contacts known in the organisations) regarding contemporaneous outbreaks in the United States of America (6), Australia (7), and Canada (51), which were unable to provide their isolates for inclusion. Although this may have revealed that multiple Genotypes were responsible for the early 2000s outbreaks around the world, subsequent studies on ST shigellosis have shown that contemporaneous outbreaks typically belong to a common Genotype across regions including across *S. flexneri 3a*, *S. flexneri 2a*, and *S. sonnei* (9) (8, 52) (49, 53), meaning it is likely that the coincident early 2000s outbreaks of sexually transmitting *S. sonnei* represented a broadly disseminated outbreak of Genotype 3.1.

### Accessory genome dynamics in early ST *S. sonnei*

Accessory genome dynamics (such as the acquisition and loss of AMR genes and virulence factors) have been previously linked with the global epidemiological success of *S. sonnei* subtypes including colicins (36), virulence factors (54) (55), and AMR (8, 9). For this reason, we explored the presence and absence of accessory genome determinants in the London and Berlin outbreaks and the reference isolates, comparing both between the city outbreaks and between the outbreak lineage and contextual isolates.

Firstly, we investigated the colicin gene profiles of the isolates as these genes may contribute to pathogen success by aiding competition with other pathogens in the microbiota of infected individuals (36). This revealed that while the 3.1 Genotype shared some colicins with other Lineages of *S. sonnei* and other Genotypes of Lineage III (Supplementary Figure 1), the acquisition of a unique colicin gene (EFY5874042) was conserved in the 3.1 Genotype relative to the other contextual isolates (**Figure 4, Supplementary Figure 1**). This is an unnamed colicin protein that reported through similarity (98.02% amino acid identity) with a colicin-like bacteriocin tRNAse domain-containing protein (WP_000012965.1) (56). Although this gene may have contributed to the early emergence of ST *S. sonnei,* the lack of its presence in subsequent outbreaks suggests this is unlikely. This is consistent with competition dynamics being common across both GBMSM and non-GBMSM host microbiota.

We then explored virulence genes across the isolates and found unique profiles across the London and Berlin outbreaks isolates which arose from both gains and losses relative to background strains (**Figure 4, Supplementary Figure 2**). There were differences in the virulence gene profiles between the two city outbreaks. Including specifically, that the Berlin outbreak isolates appear to have lost the *ygeG* and *espX1* genes (encoding a secretion-associated chaperone in the T3SS (57) and a T3SS toxic effector protein (58), respectively) and the near-unique acquisition of the *mcbA* and *trat* genes in the London outbreak (**Figure 4**). The *mcbA* gene encodes the bacteriocin microcin B17 precursor virulence protein, which participates in the killing of other bacteria (59) and the *traT* gene encodes a protein associated with an increased ability for bacteria to resist the antimicrobial effects of human serum and phagocytosis by macrophages (60) which might promote bacteraemia (61, 62). Compared with the contextual isolates however, Genotype 3.1 lacked Z2206 and ECSMS25_B0007; involved in adhesion and colicin activity respectively (63–65). However, in common with all Lineage III isolates, they had lost *hly*E, *flm*H, *flh*A, *asp*L1 and *aec*30. Finally, relative to other Lineage III isolates (but in common with Lineages I, II, and V) they retained the genes *flm*B, *flm*F, *flm*G, *eha*A, and *cfa*A (**Supplementary Figure 2**). Loss of virulence genes which encode immunogenic components has been recently demonstrated to aid in improved immune evasion (55) and these complementary findings underline that the virulence gene profile of Genotype 3.1 might represent the early evolutionary steps towards the highly globally disseminated Lineage III.

Finally, as the success of the contemporary ST *S. sonnei* (i.e. Genotypes (3.6.1.1.2 and 3.6.1.1) have been driven by their acquisition of AMR (9) (49), we explored the presence of AMR genes and potentially associated plasmids. The London and Berlin outbreak isolates had different AMR profiles compared with each other and contextual isolates (**Figure 4**). The Berlin outbreak isolates had a fully conserved (100%, *n* = 9/9) AMR profile for some AMR genes, including those associated with the integrated transposon previously associated with Lineage III (66) *sul2, sat2, dfrA1, aadA1, aph(3”)Ib, aph*(*6*)*Id, tet*(B), and most isolates also contained *bla*_TEM-1_ (67%, *n* = 6/9) and *mph*(A) (78%, *n* = 7/9) genes. One London outbreak isolate (7%, *n* = 1/14) carried *mph*(A). The presence of *mph*(A) in most of the Berlin outbreak isolates is critical, as this gene would later become a staple in MDR and XDR ST *Shigella* spp. outbreaks (8, 9), and may be indicative of early evidence for bystander resistance driven by the treatment of other sexually transmissible illnesses at the time with azithromycin (67). As *mph*(A) presence was latterly linked to the IncFII plasmids: pKSR100 and pAPR100 (13), we undertook plasmid presence / absence and plasmid mapping analyses, which revealed IncFII plasmid presence in the London outbreak isolates (**Supplementary Figure 3)**, but the presence of neither pKSR100 or pAPR100 (**Supplementary Data Table**). None (0%, *n* = 0/23) of the isolates in either outbreak carried any other genetic determinants of AMR associated with contemporary outbreaks i.e. QRDR point mutations, *bla*_CTX-M_ genes, or the *erm*(B) gene causing ciprofloxacin, ceftriaxone, and contributing to azithromycin, resistances respectively (9).

### The outbreak isolates had a most recent common ancestor in 1999

As the accessory genome dynamics of the outbreaks did not reveal any obvious events that might explain this early emergence of sexually transmissible *S. sonnei*, we conducted temporal dating of the Genotype 3.1 emergence to associate its emergence with possible host and environmental factors. Following a strong temporal signal from preliminary analyses (**Supplementary Figure 4**), we used Bayesian Evolutionary Analyses by Sampling of Trees (BEAST) to estimate the emergence date of the early 2000s outbreaks. This indicates that the most recent common ancestor first emerged in 1999 (95% HPD interval 1998 – 2000). This is consistent with the emergence date of the internationally disseminated ST *S. flexneri* 3a (1998, 95% HPD 1996 – 1998) (8). These parallel emergences coinciding is a particularly critical finding, as HAART for HIV had become readily available and accessible through the National Health Services in the UK and Germany from 1996 onwards (68, 69).

Thus, we stipulate that the time-gap between the large reporting of outbreaks of sexually transmissible shigellosis from the 1970s with those in the early 2000s may have occurred due to reduced higher-risk sexual activity during the HIV pandemic. Subsequently, the widespread availability and use of HAART for HIV from 1996 onwards (70, 71) led to increased participation in higher-risk sexual activities (68) (72) following its initial introduction and the subsequent re-emergence of ST shigellosis. This is also consistent with findings of expanding *Shigella* populations overlapping with expanding populations of people living with HIV in the HIV-hyperendemic nation of South Africa (73).

## Conclusions

Overall, this study adds greatly to our understanding of the emergence of endemically transmitting ST shigellosis. Our literature review highlighted that ST shigellosis was first detected in the early 70s, and then underwent a quiescent period until sporadic, disseminating outbreaks in the 2000s, followed by rapid spread in the 2010s associated with the acquisition of AMR. Our genomic epidemiology analyses of some of the earliest outbreaks highlight a re-emergence of ST shigellosis associated with the implementation of HAART of an early evolutionary representative of Lineage III that was poised to disseminate internationally, consistent with broader global trends in the pathogen. The relationship of this 3.1 Genotype and the broader global dissemination of Lineage III, and the full extent and role of travel more broadly in driving ST shigellosis need to be further elucidated. Future work in the area also needs to include investigation of the biological relationship between HIV and shigellosis, whether the HIV pandemic contributed to the earliest emergence, and a more intensive study of the relationship of sexual health therapeutic interventions (such as HAART, HIV pre-and post-exposure prophylaxis, and antimicrobial therapies for other sexually transmissible infections) with the development of AMR in *Shigella* and other ST enteric illnesses.

## Ethics approval and consent to participate

No ethics approved was required. No individual patient consent was required or sought as UKHSA has authority to handle patient data for public health monitoring and infection control under section 251 of the UK National Health Service Act of 2006 (previously section 60 of the Health and Social Care Act of 2001).

## Consent for publication

Not applicable. There are no names, initials, hospital numbers, images, or videos relating to an individual person present in this article or the **Supplementary Data Table**.

## Author contributions

L.C.E.M. and K.S.B. were responsible for the writing of the original draft and visualisation. All authors contributed to the review and editing of the manuscript. L.C.E.M., F.J., and K.S.B. were responsible for formal analyses, investigation, and methodology. C.J. and K.S.B. were responsible for conceptualisation. R.V, C.J, and K.S.B were responsible for funding acquisition, project administration, and supervision. A.F., C.J., and K.S.B. were responsible for data curation and providing resources.

## Data Availability

All data produced in the present work are available within the Supplementary Data Table.

## Supplementary Information

**Supplementary Figure 1.**
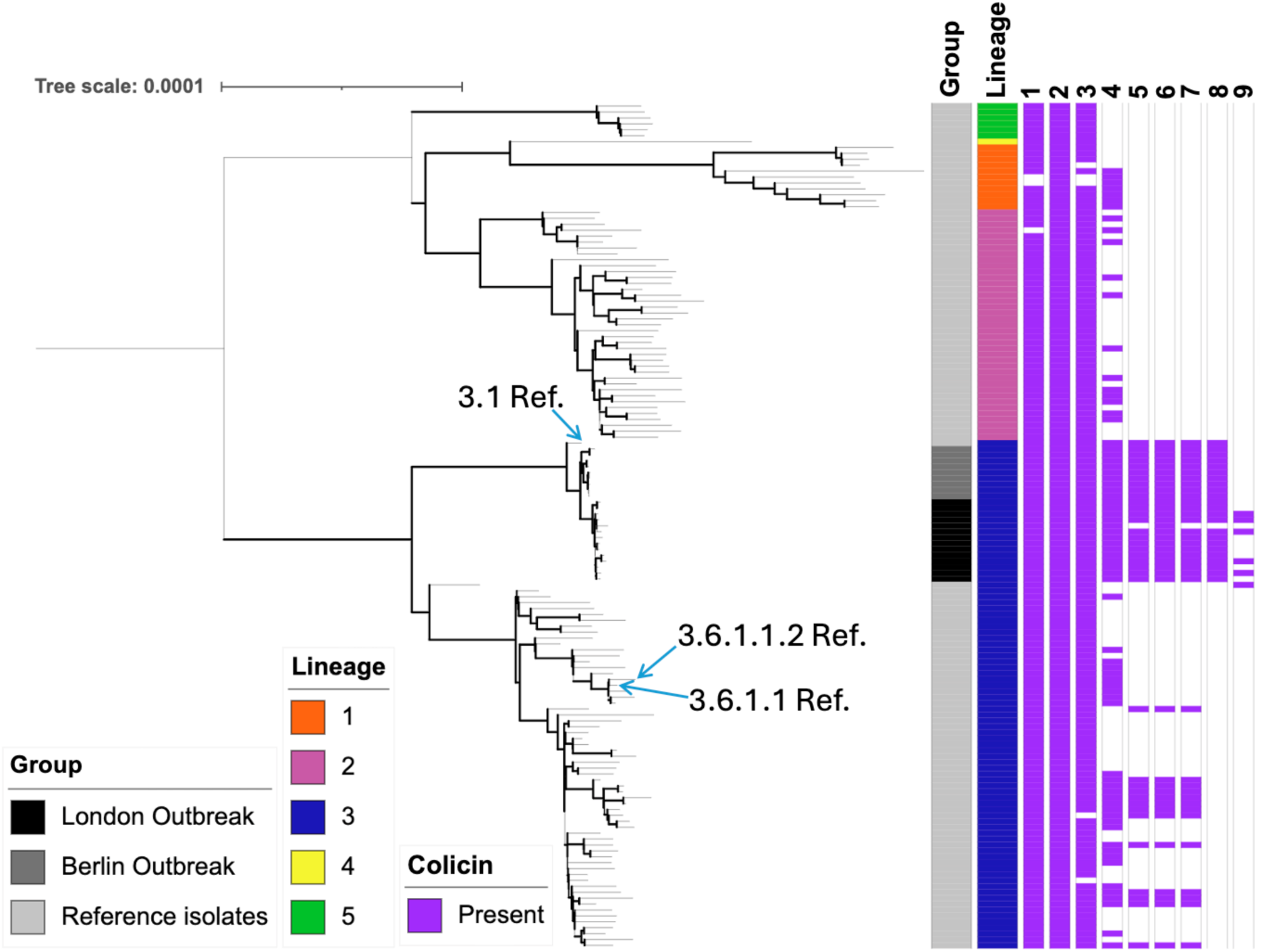
The London and Berlin outbreak isolates have a conserved colicin gene profile. The phylogenetic tree in Figure 4 with metadata tracks showing: group, lineage, and presence or absence of colicin genes which changed in proportions among groups, coloured according to the inlaid keys. Colicin genes have been represented as lane numbers, corresponding to the following: (1 = Colicin, ENA | QPR02973), (2 = Colicin Import Protein, ENA | CDU39502), (3 = Colicin-I-Receptor, ENA | EGI15668), (4 = Colicin Lysis Protein Precursor, ENA | AAN28375), (5 = Colicin T7-1, ENA | AAK67298), (6 = Colicin T7-3, ENA | CAC41014), (7 = Colicin Type 7, ENA | AAK67300), (8 = Colicin, ENA | EFY5874042), (9 = Colicin, ENA | QLM32732).

**Supplementary Figure 2.**
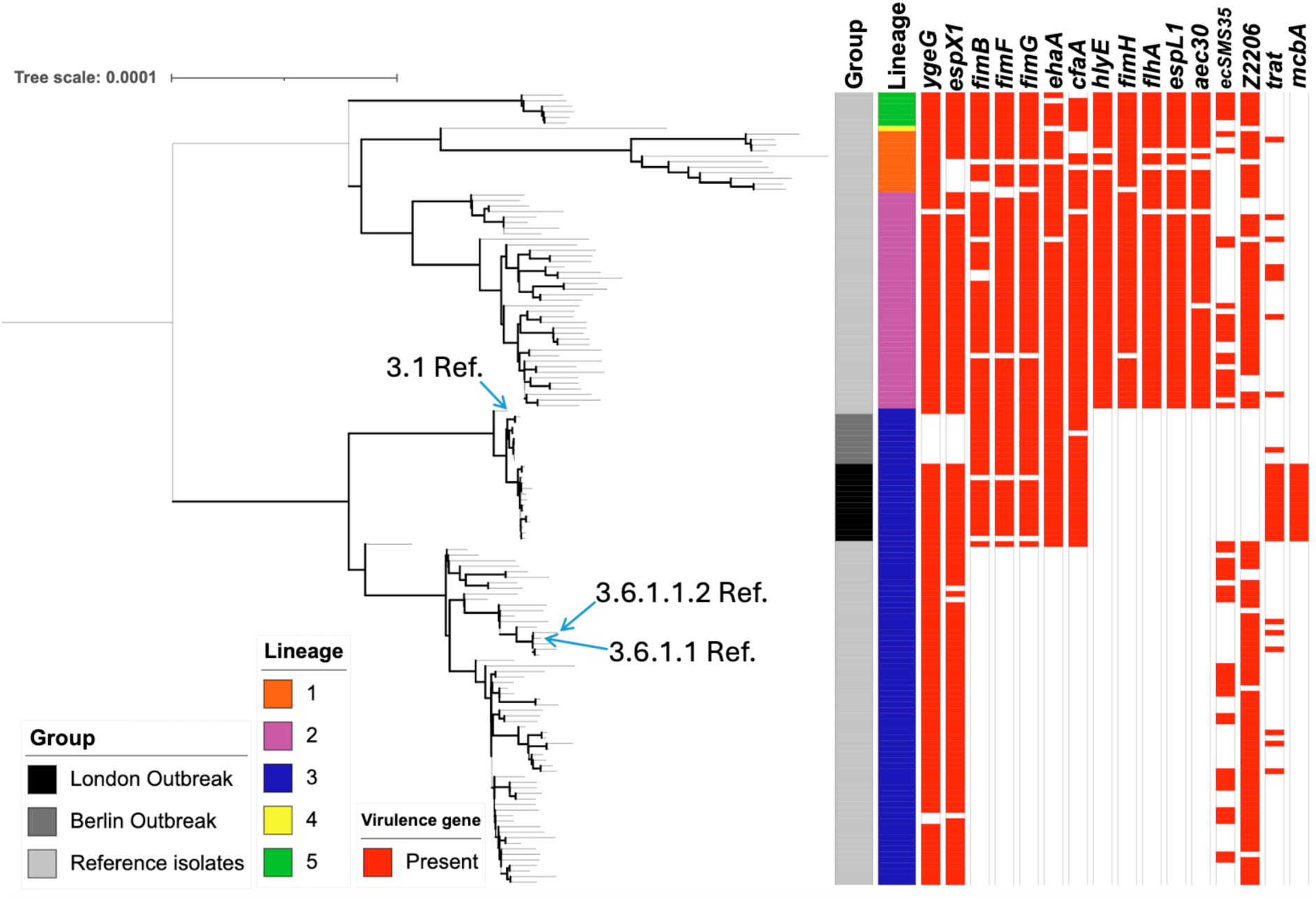
Virulence gene loss and acquisition among the London and Berlin outbreak isolates. The tree from Figure 4 is shown with metadata tracks showing, group, Lineage, and the presence or absence of selected virulence genes which change in proportion among groups of interest, coloured according to the inlaid keys.

**Supplementary Figure 3.**
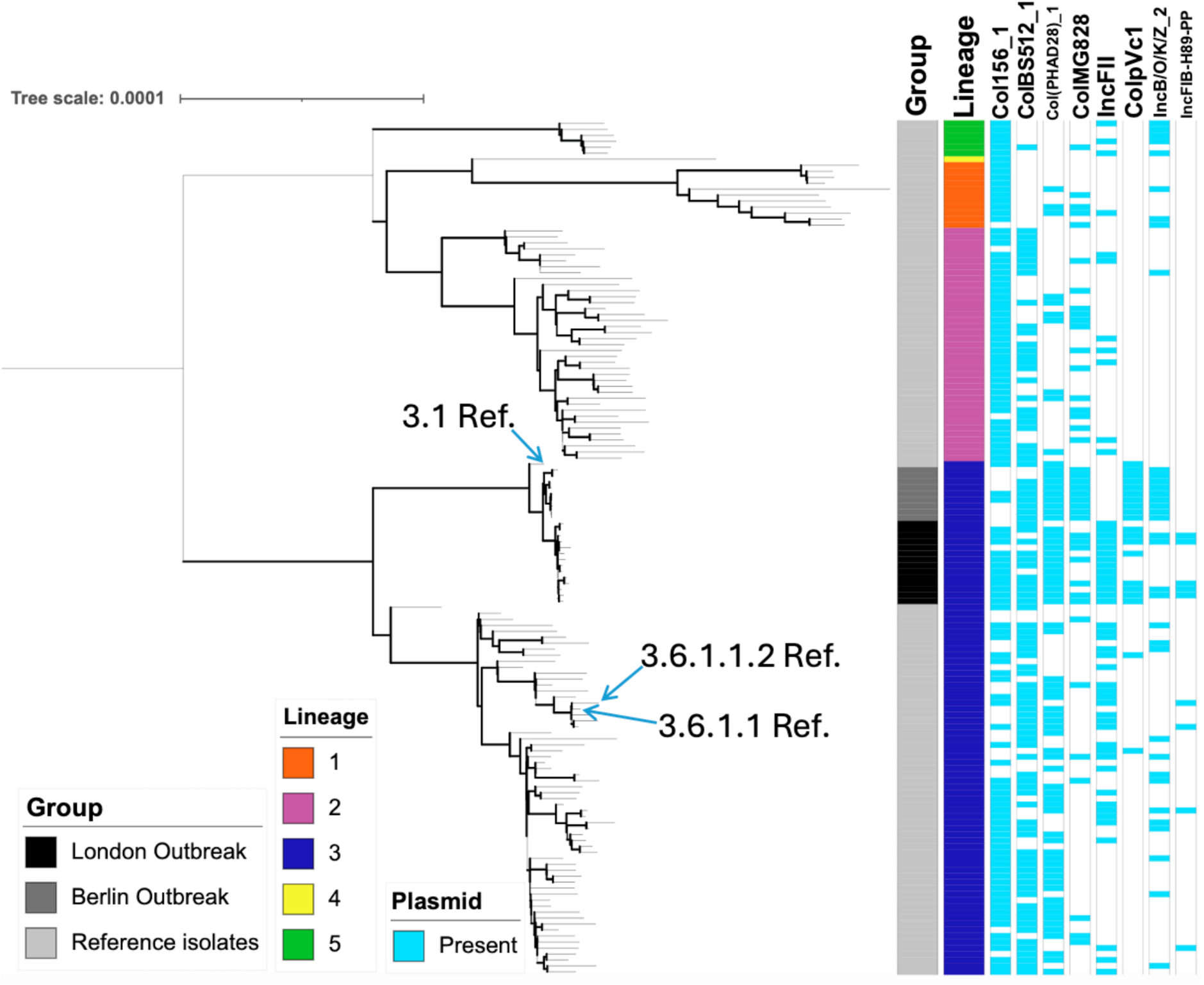
Unique plasmid conservation profiles between the London and Berlin outbreak isolates. The tree from Figure 4 is shown with metadata tracks showing, group, lineage, and presence or absence of plasmids which change in proportion among groups of interest, coloured according to the inlaid keys.

**Supplementary Figure 4.**
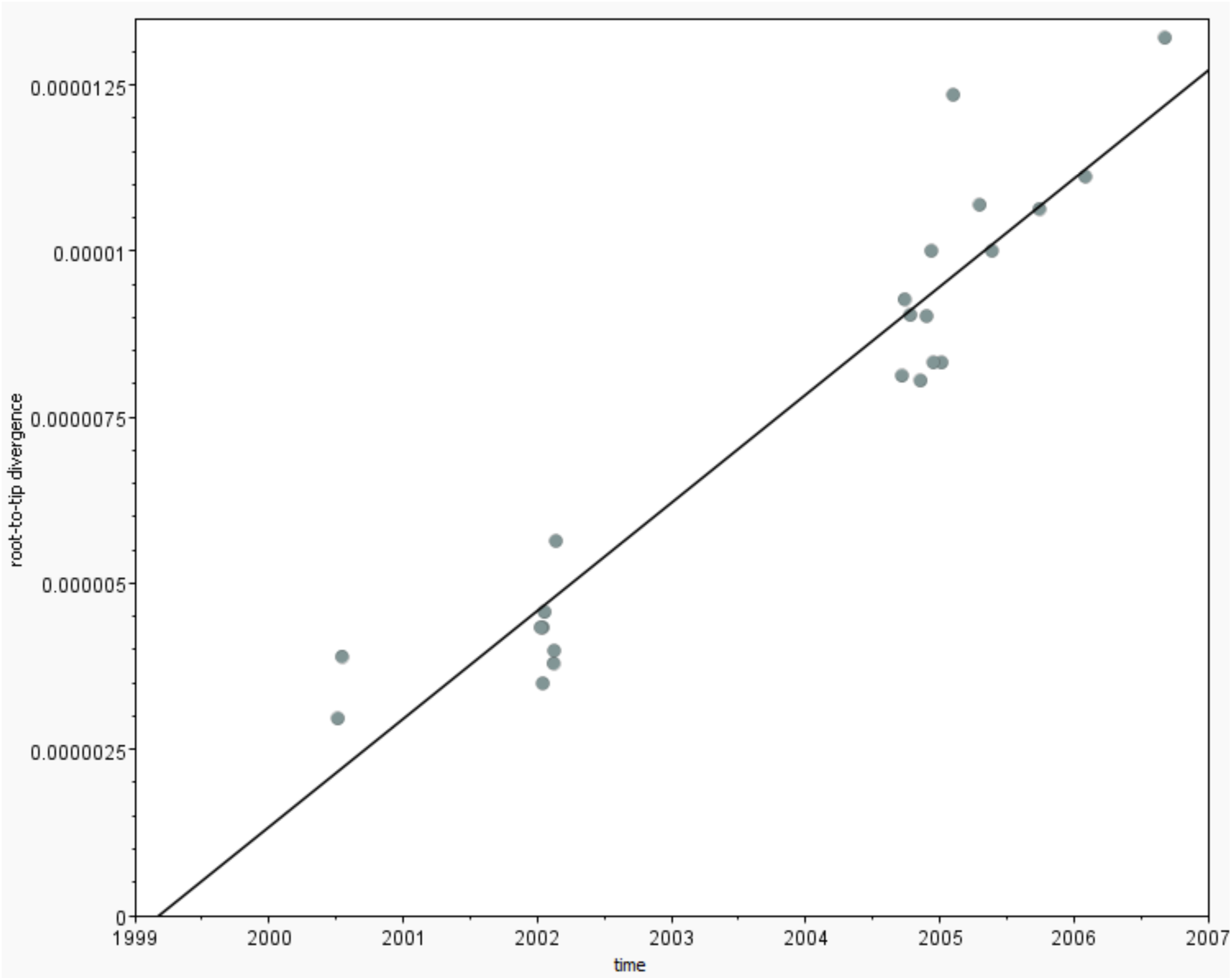
The temporal signal of the Berlin and London outbreak isolates shows most recent common ancestor evolutionary emergence as the year 1999. Root-to-Tip divergence results showing the best-fitting root, heuristic residual mean squared. Input was a Newick tree file containing the isolates of interest. Slope (rate): 1.6231E-6; X-Intercept (TMPRCA) 1999.1688; Correlation Coefficient 0.9496; R squared 0.9017, residual mean squared 1.0246E-12. The specific date (dd/MM/yyyy) of collection for all the isolates were known. Dates were specified as years since’sometime in the past’.

